# Transcriptome analyses of the cortex and white matter of focal cortical dysplasia type II: insights into disease mechanisms and tissue characterization

**DOI:** 10.1101/2022.08.23.22279011

**Authors:** Guilherme Rossi Assis-Mendonça, Maria Carolina Pedro Athié, João Vitor Gerdulli Tamanini, Arethusa de Souza, Gabriel Gerardini Zanetti, Patrícia Aline Oliveira Ribeiro de Aguiar Araújo, Enrico Ghizoni, Helder Tedeschi, Marina Koutsodontis Machado Alvim, Vanessa Simão de Almeida, Welliton de Souza, Roland Coras, Clarissa Lin Yasuda, Ingmar Blümcke, André Schwambach Vieira, Fernando Cendes, Iscia Lopes-Cendes, Fabio Rogerio

## Abstract

Focal cortical dysplasia (FCD) is a common cause of pharmacoresistant epilepsy. According to the 2022 International League Against Epilepsy classification, FCD type II is characterized by dysmorphic neurons (IIa and IIb) and may be associated with balloon cells (IIb). We present a multicentric study to evaluate the transcriptomes of the gray and white matters of surgical FCD type II specimens. We aimed to contribute to pathophysiology and tissue characterization. We investigated FCD II (a and b) and control samples by performing RNA-sequencing followed by immunohistochemical validation employing digital analyses. We found 342 and 399 transcripts differentially expressed in the gray matter of IIa and IIb lesions compared to controls, respectively. The top enriched cellular pathway in IIa and IIb gray matter was cholesterol biosynthesis, the genes *HMGCS1, HMGCR*, and *SQLE* being upregulated in both type II groups. We also found 12 differentially expressed genes when comparing transcriptomes of IIa and IIb lesions. Only 1 transcript (*MTRNR2L12*) was significantly upregulated in FCD IIa. The white matter in IIa and IIb lesions showed 2 and 24 transcripts differentially expressed, respectively, compared to controls. No enriched cellular pathways were detected. *GPNMB*, not previously described in FCD samples, was upregulated in IIb compared to IIa and control groups. Upregulations of cholesterol biosynthesis enzymes and *GPNMB* genes in FCD groups were immunohistochemically validated. Such enzymes were mainly detected in both dysmorphic and normal neurons, whereas GPNMB was observed only in balloon cells. Overall, our study contributed to identifying cortical enrichment of cholesterol biosynthesis in FCD type II, which may correspond to a neuroprotective response to seizures. Moreover, specific analyses in either the gay or the white matter revealed upregulations of *MTRNR2L12* and GPNMB, which might be potential neuropathological biomarkers of a cortex chronically exposed to seizures and of balloon cells, respectively.

## 1. Introduction

Epilepsy is a neurological disease characterized by recurrent unprovoked seizures, which may be caused by different brain disorders. Most patients have a satisfactory clinical outcome with appropriate medications. However, seizures may be pharmacoresistant and have detrimental and even life-threatening consequences (1,2). Around 30% of the patients with focal epilepsies are candidates for surgical treatment, and their clinical outcome depends on the pathological condition causing the seizures (3)The commonest causes leading to medically intractable epilepsy are hippocampal sclerosis, long-term epilepsy-associated brain tumors, and malformation of cortical development (MCD) (4). Particularly, MCDs represent a wide range of lesions arising from the disruption of steps of cortical formation: cell proliferation, cell migration, and cortical organization. Although the pathogenesis is still unclear, specific genetic defects have been identified (5–7).

The incidence of MCDs in individuals submitted to epilepsy surgery varies among centers. However, focal cortical dysplasia (FCD) is the most frequently reported MCD. FCD is a structural lesion with different sizes, locations, and histopathological findings (7). Clinically, seizure features depend on the FCD localization, and electroencephalography mostly shows epileptiform discharges spatially correlated with the lesion. On magnetic resonance imaging (MRI), FCDs may present with cortical thickening associated with cortical–white matter junction blurring, a hyperintense signal on T2-weighted images, and an abnormal pattern of sulci and gyri (1,4). Historically, the neuropathological classification of FCD has been as variable as its broad morphological spectrum (4,7–9). Recently, the International League Against Epilepsy (ILAE) has reviewed and updated its neuropathological classification system for FCD. Specifically, FCD type II refers to isolated lesions characterized by dyslamination of the cortex and dysmorphic neurons without (IIa) or with balloon cells (IIb) (10,11).

The ILAE classification was expected to improve comparisons between electroclinical, imaging and/or seizure control studies from different institutions and data obtained from molecular investigations on the pathophysiology of FCDs (12). In this context, we designed an original study in which transcriptome analysis (12) was performed to identify differentially expressed mRNAs in the cortex and white matter of individuals with epilepsy due to FCD type IIa or IIb. We compared the transcriptome profiles of the two lesions and those obtained from control specimens from autopsied individuals without a history of neurological disease. Immunohistochemical validation was performed for selected molecules. We aimed to improve the current understanding of tissue findings associated with chronic seizures and find potential FCD type II biomarkers by using RNA-Seq transcriptome analysis to simultaneously evaluate a wide range of molecular pathways.

## 2. Materials and Methods

### 2.1. Surgical samples and groups

This is a retrospective study for which the samples were obtained in biorepositories from both Institutions (University of Campinas, Brazil, and University Hospital Erlangen, Germany). We obtained these samples from patients without the restriction of age or gender but with previous confirmation that the surgical procedure involved the brain area with clinically and MRI determined FCD. We evaluated only frontal lobes for transcriptome analyses, as this is the most common location for FCD ILAE type II (13). We studied additional specimens from other brain regions with histopathologic confirmation of FCD ILAE type II for immunohistochemical validation. We obtained control samples from frontal lobes of adults submitted to autopsy (between 6 and 12 hours *post mortem*) with neither neurological disease nor a history of seizures. Details on clinical information and allocation of surgical and control specimens for molecular and histopathological analyses are presented in Tables 1 and 2.

### 2.2. Brain sampling and neuropathological diagnoses

Fresh brain samples were collected from surgical and autopsy specimens and (i) formalin-fixed and paraffin-embedded (FFPE) or (ii) snap frozen in liquid nitrogen and stored at -80°C.

FFPE samples were submitted to diagnostic routine, that is, evaluation of the cortical cytoarchitecture and cellularity and myelination of the white matter in serial 4 μm-sections stained with hematoxylin and eosin (H&E) and submitted to immunohistochemical reactions. For the latter protocol, the sections were exposed to antibodies against NeuN (neuronal marker; 1:1000, clone A60, Merck Millipore, cat#MAB377, Temecula, CA, USA), MAP2 (neuronal marker; 1:1000, clone M13, Thermo Fisher, cat#13-1500, Waltham, MA, USA), SMI 32 (neuronal marker; 1:2500, clone SMI 32, Biolegend, cat#SMI-32R, San Diego, CA, USA), GFAP (astrocytic marker, 1:100, clone 6F2, Dako/Agilent, cat#M0761, Santa Clara, CA, USA), vimentin (1:100, clone V9, e-Bioscience/Thermo Fisher, cat#14-9897-82, Waltham, MA, USA), CD34 (1:50, clone QBEnd-10, Dako, cat#M7165, Glostrup, Denmark) and CNPase (myeloarchitecture marker; 1:500, clone 11-5 B, Millipore, cat#MAB326, Darmstadt, Germany), for 18h at 4°C. Then, a detection solution containing the secondary antibody and peroxidase (AdvanceTMHRP®, Dako, cat#K4068, Glostrup, Denmark; or EnvisionTM Flex+, Dako, cat#K8002, Glostrup, Denmark) was added for 30 min at 37°C. 3,3-diaminobenzidine (DAB) was used as chromogenic substrate and counterstaining was performed with hematoxylin. Negative controls (without primary antibody) were run concurrently with all immunohistochemical reactions.

Samples exhibiting cortical dyslamination, hypertrophic and dysmorphic neurons (disoriented neurons with anomalous cytoplasmic distribution of Nissl substance and accumulation of non-phosphorylated neurofilament (SMI 32-positive)) without or with balloon cells (large cells with opaque eosinophilic cytoplasm, vesicular nucleus and immunopositivity for vimentin) were classified as FCD ILAE type IIa or IIb, respectively (10, 13). Samples from autopsies (control group) were submitted to the same protocol to exclude microscopic alterations. All histological analyses were performed at regions of gyri perpendicularly cut to the pial surface, as recommended by the FCD ILAE Classification (10, 13).

### 2.3. Isolation of cortex and white matter and transcriptome analysis

Frozen samples corresponding to previously evaluated FFPE sections were serially cut (40 μm), mounted in PEN membrane covered slides (Life Technologies), immediately stained with Cresyl Violet and dehydrated with an ethanol series. The gray (cortical layer) and white matters were mechanically dissected from 3 – 4 sections per sample with the aid of a scalpel, collected in RNAse-free individual tubes and stored at -80°C.

For cDNA library preparation and next-generation sequencing, total RNA from each dissected sample was extracted and purified with Trizol (Thermo Fisher Scientific, cat # 15596018, Waltham, MA, USA). cDNA libraries were produced from extracted RNA (200 ng) by using the TruSeq Stranded total RNA kit (Illumina, San Diego, CA, USA) and sequenced in a HiSeq 2500 platform (Illumina, San Diego, CA, USA) in High-Output mode, producing 100-bp paired-end sequences. The amount (total number and number of sequences produced per sample) and quality (% of bases over Q30) of the generated sequences were also assessed. Sequence alignment was performed with STAR (https://github.com/alexdobin/STAR) to *Homo sapiens* GRCh37/hg19 assembly.

The DESeq2 (http://www.bioconductor.org/packages/release/bioc/html/DESeq2.html) package was used for transcriptome analyses. A list of differentially expressed genes with a statistical significance set at a p < 0.05 (after correction for multiple tests, *i*.*e*. adjusted p-value) was generated. Such list was used for gene ontology analysis (calculation of enrichment of pathways for the set of differentially expressed genes) by using Enrichr’s web-based tools (https://amp.pharm.mssm.edu/Enrichr/) (14,15). The results regarding the expression of genes of interest were presented as Fold change, a simplified designation that refers to the log_2_Fold change calculation performed by the DESeq2 package when comparing the different groups with each other.

### 2.4. Sequencing Analysis of mTOR Pathway Genes

We investigated the presence of somatic mutations in 68 genes of the mTOR pathway in two specimens from which a blood sample was also available (11). We used genomic DNA extracted from brain tissue surgically resected and compared it to sequences obtained from the blood samples. We used the Sure Select Human All Exon V6 ® (Agilent, Santa Clara, USA), followed by sequencing in a Hiseq 2500® (Illumina, San Diego, USA). Mosaicism was evaluated by using Mutect2.20 (16) and Strelka2 (17). Only variants identified in both programs were recorded. Somatic mosaicism was identified when <10% of reads were not aligned to the human genome (GRCh37/hg19) and were present only in surgically removed tissue sample. The impact of variants on protein function was analyzed by using SIFT, Polyphen2, FATHMM, and MutationTaster (http://sift.jcvi.org; http://genetics.bwh.harvard.edu/pph2/; http://fathmm.biocompute.org.uk; http://www.mutationtaster.org). The classification recommended by the American College of Medical Genetics and Genomics (18) was used to interpret sequence variants as implemented in the Franklin™ software (GNX Data Systems, Inc., https://franklin.genoox.com/clinical-db/home). We searched the variants identified in two control databases: GnomAD (https://gnomad.broadinstitute.org) and BipMed (www.bipmed.org) (19).

### 2.5. Immunohistochemical detection of proteins encoded by differentially expressed genes

For validation of the transcriptome results, 4μm-thick sections from FFPE specimens were submitted to immunohistochemical reactions. The sections were incubated with primary antibodies against HMGCS1, HMGCR, SQLE and GPNMB (please refer to item 3.4) for 18 h at 4°C. Specifications of each antibody are as follows: HMGCS1 (1:500, polyclonal, Abcam, cat# ab155787, Cambridge, United Kingdom), HMGCR (1:300, clone CL0260, Abcam, cat# ab242315, Cambridge, United Kingdom), SQLE (1:300, polyclonal, Sigma-Aldrich/Merck, cat#HPA020762, Darmstadt, Germany) and GPNMB (1:100, clone SP299, Abcam, cat# ab227695, Cambridge, United Kingdom). Afterwards, the detection system containing secondary antibody and peroxidase (AdvanceTMHRP®, Dako, cat # K4068, Glostrup, Denmark) was added for 30 min at 37 °C. DAB was used as chromogenic substrate and hematoxylin as counterstain. Negative controls (without primary antibody) were run concurrently with all reactions.

Following the immunohistochemical reactions, all slides were scanned by using an Aperio Scanscope CS2 (#23CS100) device. Ten representative pictures of each slide, at a 200x magnification, were chosen for the quantification of each marker, which was performed in the ImageJ® software environment. The IHC Profiler plugin (20) was applied to each image in the deconvolution step, that is, separation of hematoxylin and DAB channels. The corresponding DAB channel picture was used for quantification via the “threshold” tool. A threshold value was determined so that the histological findings in each picture were preserved in the most accurate way considering the original photo. The final image consisted of a black-and-white picture from which the percentage of positive pixels was obtained.

The Kruskal-Wallis test and the post hoc Dunn tests were performed with the GraphPad Prism software (version 8.0) for statistical analyses. Statistical significance was set at a p < 0.05.

## 3. Results

### 3.1. Clinical and neuropathological data

Seventeen FCD specimens from the UNICAMP (Brazil) and FAU (Germany) biorepositories were assessed in the transcriptome study. Patients whose samples were histopathologically assigned as FCD ILAE type IIa (n=9) or FCD ILAE type IIb (n=8) were submitted to presurgical evaluation consisting of routine EEGs, video-EEG monitoring, neuropsychological assessment, magnetic resonance imaging (MRI), and Fluorodeoxyglucose-Positron Emission Tomography (FDG-PET) and ictal Single-Photon Emission Computed Tomography (SPECT) studies when indicated. Information on the number of individuals per group and clinical data regarding gender, age at epilepsy onset, age at surgery and epilepsy duration is shown in Table 1. All patients presented MRI features of FCD type II that included focal cortical thickening, a varying degree of increased cortical and subcortical signal intensity in T2-weighted and Fluid Attenuated Inversion Recovery (FLAIR) sequences, blurring of the gray-white matter junction, focal abnormal cortical gyration, and cerebrospinal fluid cleft-cortical dimple (Fig. 1). The hyperintense T2-FLAIR signal in the subcortical white matter with a wedge shape extending to the ipsilateral ventricle ependymal surface (transmantle sign) was present in some but not all patients with FCD type IIb.

**Fig. 1.**
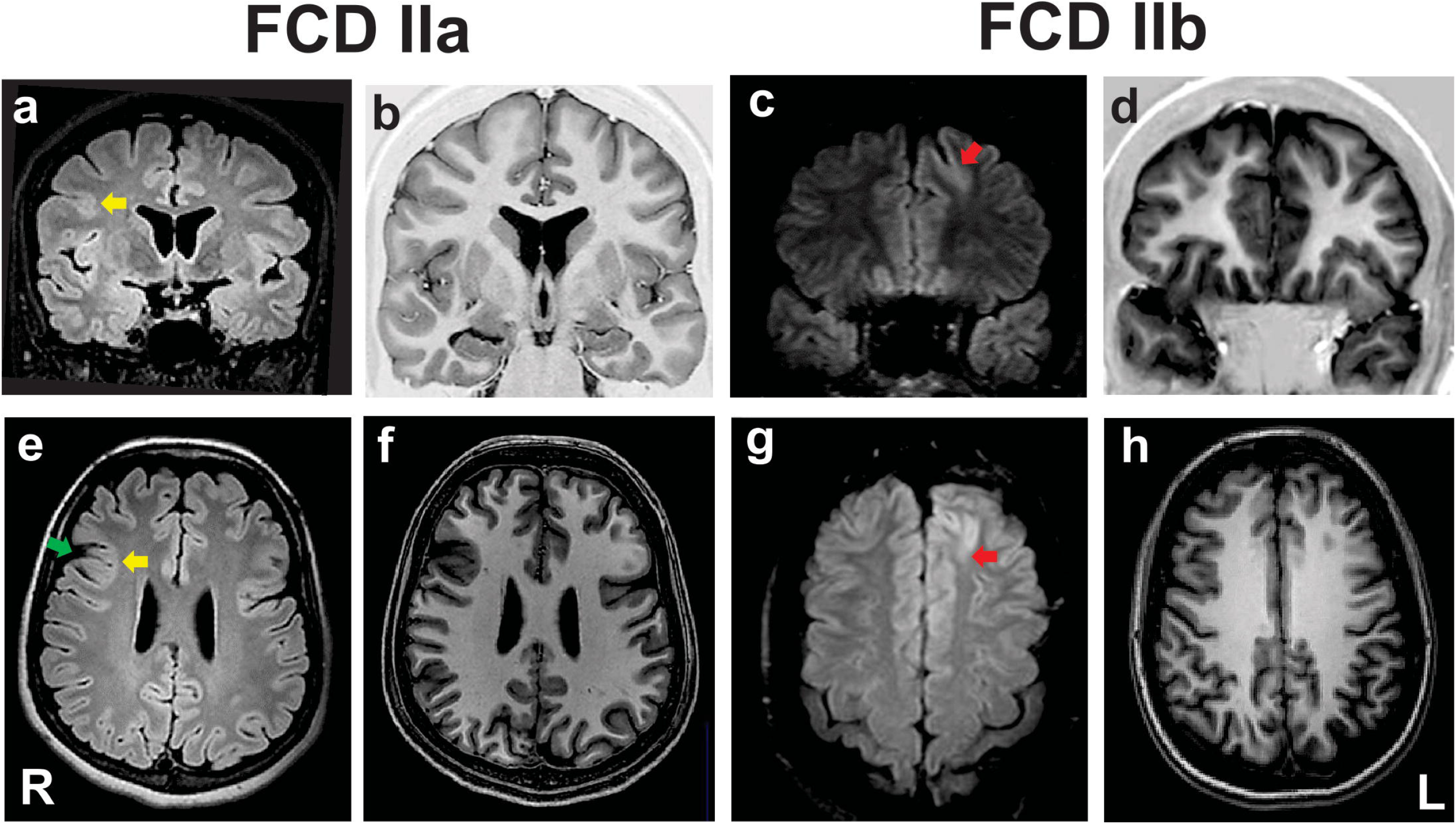
Preoperative coronal fluid-attenuated inversion recovery (FLAIR; a and c) and T1 inversion recovery (b and d), and axial FLAIR (e and g) and T1-weighted images (f and h) of two patients with pharmacoresistant frontal lobe epilepsy whose surgical samples showed focal cortical dysplasia (FCD) ILAE type IIa (a, b, e and f) and FCD type IIb (c, d, g and h). In the patient with FCD IIa there is a discrete hyperintense signal of gray matter in T2/FLAIR (yellow arrows in a and e), mild blurring of the gray-white matter junction in T1 scans, and a cortical dimple indicated by the green arrow in e. The patient with FCD type IIb has a deep sulcus with a mildly thickened cortex and hyperintense FLAIR signal in the left frontal lobe (red arrows in c and g). ILAE: International League Against Epilepsy; L: left side; R: right side

The neuropathological evaluation revealed samples with either cortical dyslamination and dysmorphic neurons (FCD IIa diagnosis) or cortical dyslamination, dysmorphic neurons and balloon cells (FCD IIb diagnosis). Gliosis and gray-white matter blurring in both FCD types were shown by increased GFAP positivity and irregularly distributed CNPase immunostaining, respectively. Heterotopic and dysmorphic neurons, as well as balloon cells, were also observed in the white matter of FCD II samples (Fig. 2). Moreover, immature CD34-positive cells were detected neither in the gray nor in the white matter.

**Fig. 2.**
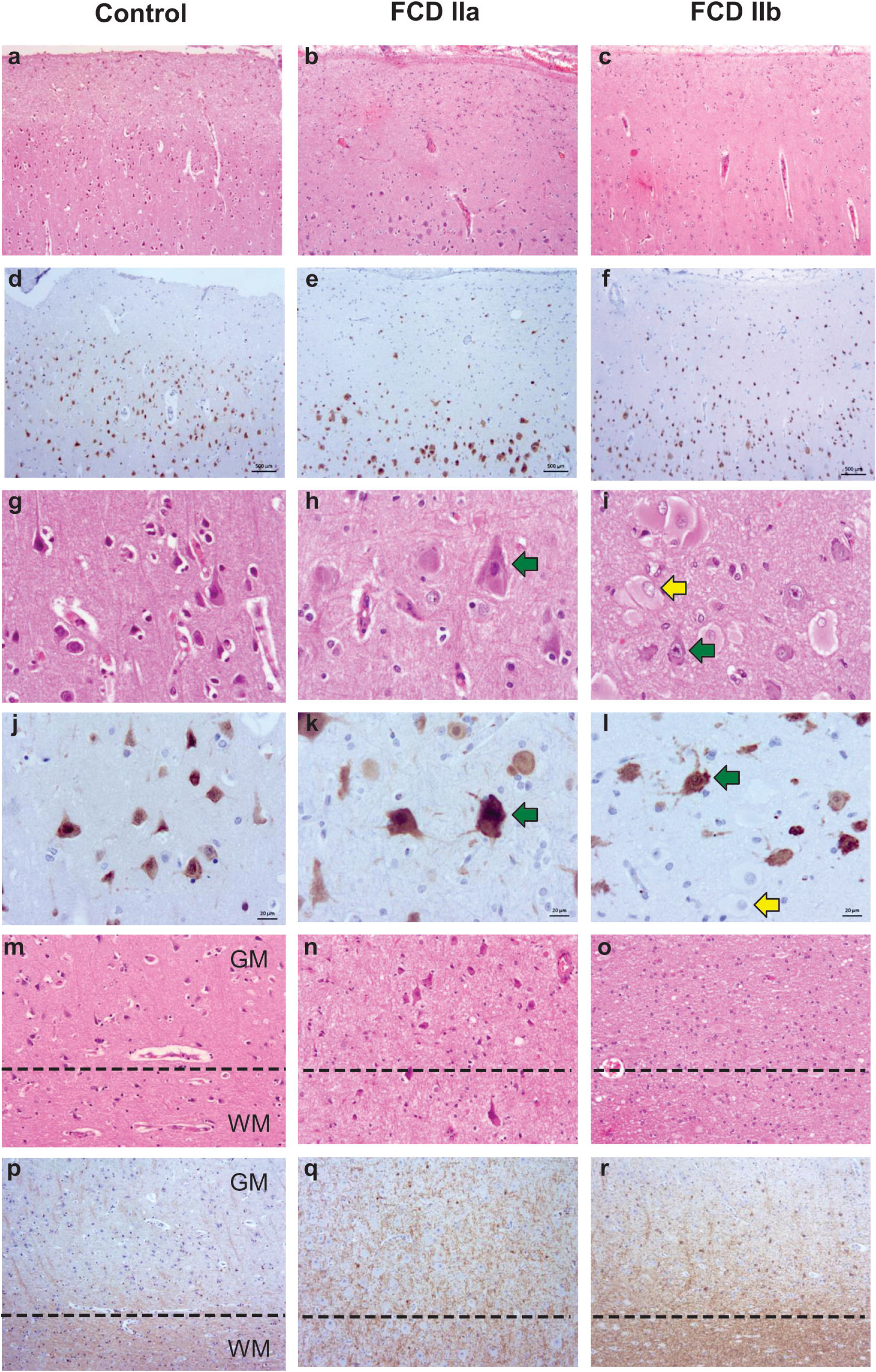
Representative histopathological and immunostaining features of autopsy (control), focal cortical dysplasia (FCD) ILAE type IIa (FCD IIa) and FCD ILAE type IIb (FCD IIb) samples. Hematoxylin and eosin staining (a-c; g-i; m-o). Immunohistochemistry for NeuN (d-f; j-l) or CNPase (p-r). Cortical cytoarchitectural features, that is, layering and neuronal size and orientation, are preserved in controls (a, d, g, j, m) specimens. However, dyslamination (b, c, e, f, h, i, k, l, n, o) and dysmorphic neurons (green arrows in h, i, k, l) without and with balloon cells (yellow arrows in i and l) are observed in FCD IIa and FCD IIb, respectively. Gray matter (GM) and white matter (WM) boundary (dotted line in m-o and p-r) is preserved in control samples (m and p) and blurred in FCD IIa (n and q) and FCD IIb (o and r). Scale bars: 500 μm (a-f), 20 μm (g-l) and 50 μm (m-r)

Control specimens (n=4 for transcriptome analyses) were obtained from autopsied individuals without a history of neurological diseases (clinical information is presented in Table 2). Histological evaluation showed neither cortical dyslamination nor cytological abnormalities in the gray and white matter. (Fig. 2).

### 3.2. Transcriptome analysis

Brazilian and German samples classified as FCD IIa were considered as one group for gene expression analyses, the same for FCD IIb specimens. Regarding the gray matter (cortical layer), a total of 342 and 399 differentially expressed genes were identified after comparing FCD IIa and FCD IIb to controls, respectively. In the comparison between FCD IIa and FCD IIb samples, we found 12 genes differentially expressed. Moreover, the principal component analysis (PCA) dimensionality reduction method based on the RNA-Seq data demonstrated that FCD IIa and FCD IIb samples aggregated in distinct clusters (Fig. 3).

**Fig. 3.**
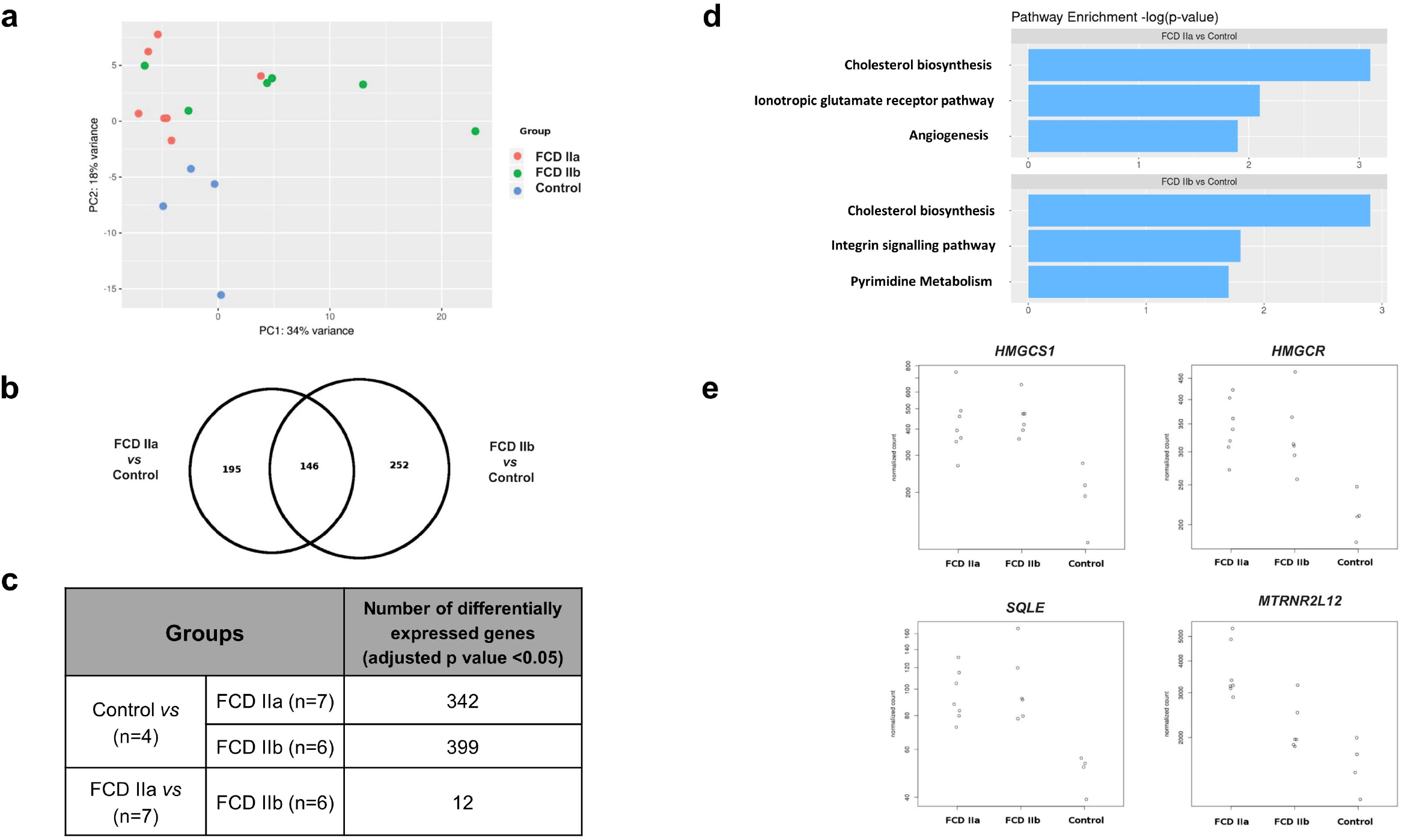
Gene expression analyses of the gray matter (cortical layer) of autopsy (control), focal cortical dysplasia (FCD) ILAE type IIa (FCD IIa) and FCD ILAE type IIb (FCD IIb) samples. a Graphical presentation of gene expression data after analyses performed with the PCA dimensionality reduction method. Brazilian and German FCD specimens with the same histopathological classification are shown as one group (FCD IIa or FCD IIb). Note that the majority of FCD IIa samples tended to group together, as observed for FCD IIb cases. Each circle corresponds to an individual sample. b Venn diagram showing the number of differentially expressed genes in each group compared to controls. FCD IIa and FCD IIb groups showed 342 and 399 differentially expressed genes, respectively. A total of 146 genes were common to FCD IIa and IIb groups. The highest number of exclusively differentially regulated genes was verified in FCD IIb (252). c Number of differentially expressed genes when comparing the groups. d Graphs depicting the three main enriched pathways containing the highest numbers of differentially expressed genes represented in the Venn diagram for FCD IIa and FCD IIb (*vs* control group). Values presented in the X-axis correspond to Enrichr -log(p-value) by using Panther Pathways 2016 database. Pathways were considered statistically significant when associated with values higher than -log(0.05) = 1.30. Additional data on enriched pathways and details on gene expression are reported in the Supplementary Material. e Expression of the differentially expressed genes *SQLE, HMGCS1, HMGCR* and *MTRNR2L12* in each group. Please refer to the text for details on fold change for each gene and the corresponding statistical significance (adjusted p-value).

An enrichment pathway analysis was performed for the differentially expressed genes in FCD IIa or FCD IIb *versus* the control group by using the Enrichr tool. The main pathways found considering the Panther 2016 database were Cholesterol biosynthesis, Ionotropic glutamate receptor pathway, and Angiogenesis for FCD IIa. In FCD IIb, the main enriched pathways were Cholesterol biosynthesis, Integrin signaling pathway, and Pyrimidine metabolism (Fig. 3; enriched pathways and the corresponding differentially expressed genes are presented as Supplementary Material).

When taking into account genes less frequently or not previously investigated in human FCDs, we found upregulation of 3 cholesterol biosynthesis genes in FCDs, namely *HMGCS1* (3-Hydroxy-3-Methylglutaryl-CoA Synthase 1; FCD IIa *vs* Control: Fold change = 0.92, adjusted p-value= 0.001; FCD IIb *vs* Control: Fold change = 0.98, adjusted p-value= 0.001); *HMGCR* (3-Hydroxy-3-Methylglutaryl-CoA Reductase; FCD IIa *vs* Control: Fold change = 0.63, adjusted p-value= 0.005; FCD IIb *vs* Control: Fold change = 0.59, adjusted p-value= 0.01); and *SQLE* (Squalene Epoxidase; FCD IIa *vs* Control: Fold change = 0.76, adjusted p-value= 0.01; FCD IIb *vs* Control: Fold change = 0.89, adjusted p-value= 0.002) (Fig. 3).

Moreover, a subsequent analysis focusing on the differentially expressed genes between FCD IIa and FCD IIb showed that *MTRNR2L12* was the only upregulated transcript in FCD IIa (FCD IIa *vs* Control: Fold change = 1.09, adjusted p-value = 0.0000003; FCD IIa *vs* FCD IIb: Fold change = 0.67; adjusted p-value=0.05) (Fig. 3).

As regards the white matter, the comparison of FCD IIa and FCD IIb with the control group showed a total of 2 and 24 differentially expressed genes, respectively. When comparing FCD IIa and FCD IIb groups, we found 4 genes differentially expressed (Fig. 4; Supplementary Material). No clustering of samples was identified by using the PCA method and the corresponding RNA-Seq data. Similarly, the Enrichr tool found no enriched pathway considering the three groups and the Panther 2016 database. Also, we performed analyses focusing on differentially expressed genes not previously reported in human FCDs. Particularly, *GPNMB* (glycoprotein nonmetastatic melanoma protein B) was upregulated in FCD IIb *vs* Control (Fold change = 4.50, adjusted p-value = 0.03) and downregulated in FCD IIa *vs* FCD IIb (Fold change = -5.68, adjusted p-value = 0.0000124) (Fig. 4).

**Fig. 4.**
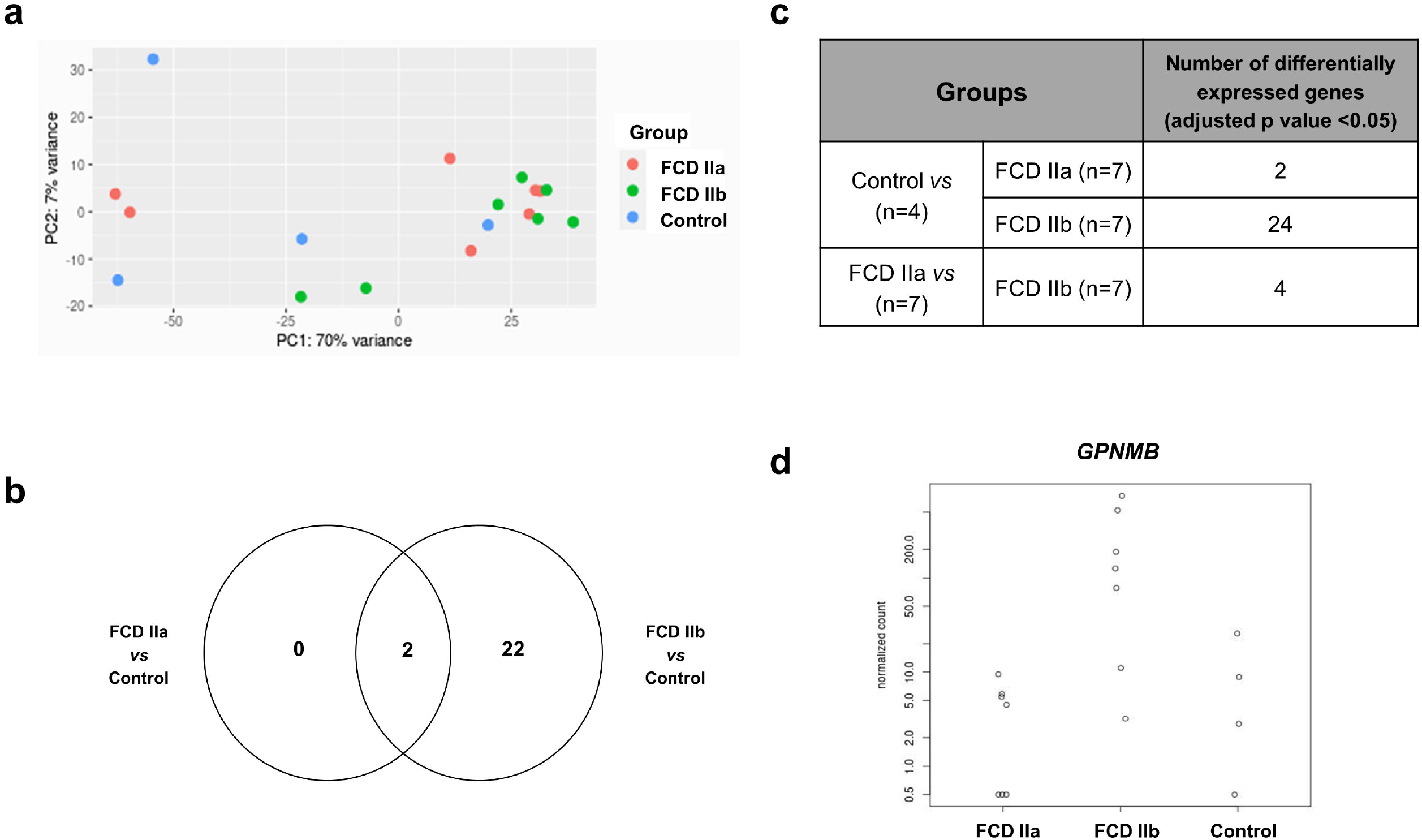
Gene expression analyses of the white matter of autopsy (control), focal cortical dysplasia (FCD) ILAE type IIa (FCD IIa) and FCD ILAE type IIb (FCD IIb) groups. a Graphical presentation of gene expression data after analyses performed with the PCA dimensionality reduction method. Brazilian and German FCD specimens with the same histopathological classification are shown as one group (FCD IIa or FCD IIb). Samples with the same neuropathological diagnosis did not cluster together. b Venn diagram showing the number of differentially expressed genes in each group compared to controls. FCD IIa and FCD IIb groups showed 2 and 24 differentially expressed genes, respectively. Two genes were common to FCD IIa and IIb groups. The highest number of exclusively differentially regulated genes was verified in FCD IIb (22). c Number of differentially expressed genes when comparing the groups. d Expression of the differentially expressed gene *GPNMB* in each group. Please refer to the text for details on fold change and the corresponding statistical significance (adjusted p-value). Additional data on gene expression are reported in the Supplementary Material.

### 3.3. Sequencing Analysis of mTOR Pathway Genes

This additional characterization refers to Individuals #13 and #14 with FCD type IIb presented in Table 1. We found a variant classified as likely pathogenic in brain tissue resected by surgery in one sample: gene *MTOR*, c.5930C>G, p.Thr1977Arg, 3% in tissue resected by surgery, and 0% in blood (Individual #13). This variant has not been described in control databases; however, it has been reported as a somatic mutation in tissue with focal cortical dysplasia (FCD) (21) In addition, we found the variant c.4375G>C, p.Ala1459Pro, 2.4% in tissue resected by surgery and 0% in blood, classified as a variant of uncertain significance (VUS) (Individual #14). This variant has not been reported in control databases or samples with FCD.

### 3.4. Immunohistochemical validation of the transcriptome results

In this step, we used a total of 24 samples from Brazilian individuals (6 FCD IIa, 10 FCD IIb, and 8 controls; Tables 1 and 2). Considering the differential expressions of *HMGCS1, HMGCR, SQLE* and *GPNMB* in FCD groups, we further evaluated immunohistochemically the expression of the codified proteins by using available commercial antibodies.

#### 3.4.1. HMGCS1, HMGCR and SQLE

Immunostaining for the cholesterol biosynthesis enzymes HMGCS1, HMGCR and SQLE showed a diffuse cytoplasmic pattern, mainly observed in normal and dysmorphic neurons. Particularly, in FCD groups, immunopositive abnormal neurons were more frequently observed than their normal counterparts. Balloon cells were virtually immunonegative (Fig. 5).

**Fig. 5.**
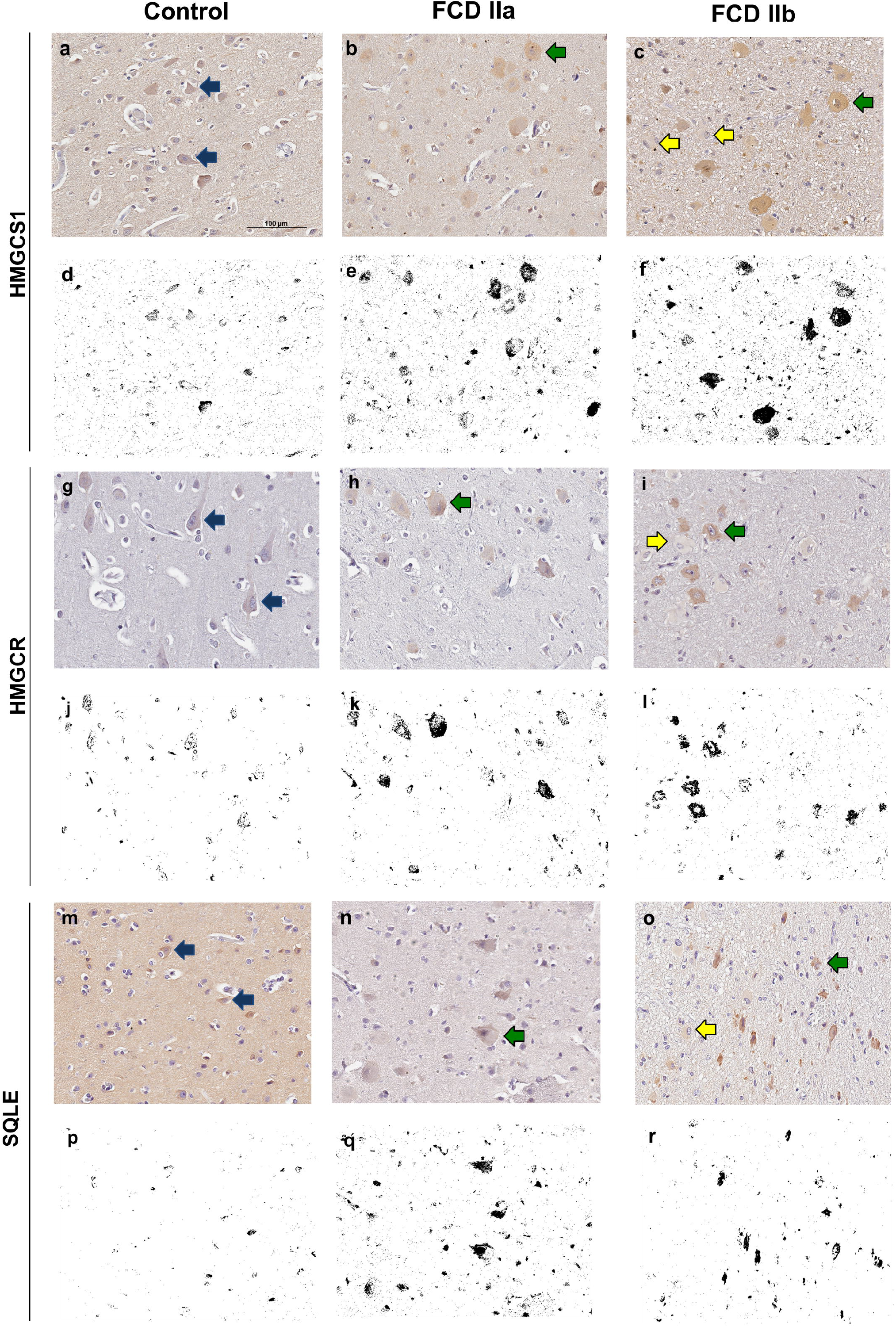
Immunohistochemical reactions for 3-Hydroxy-3-Methylglutaryl-CoA Synthase 1 (HMGCS1), 3-Hydroxy-3-Methylglutaryl-CoA Reductase (HMGCR) and Squalene Epoxidase (SQLE) in the cortical layer of control (a, g, m), focal cortical dysplasia (FCD) ILAE type IIa (FCD IIa) (b, h, n) and FCD ILAE type IIb (FCD IIb) (c, i, o) specimens. In FCD IIa and FCD IIb, a cytoplasmic staining was observed in dysmorphic neurons (green arrows in b, c, h, i, n and o), in contrast with the negativity in balloon cells of FCD IIb lesions (yellow arrows in c, i and o). Normal neurons (blue arrows in a, g, m) showed less noticeable cytoplasmic positivity compared to the dysmorphic neurons. Digital images of the presented immunohistochemical reactions were obtained by using the ImageJ® software (d-f, j-l, p-r). In this setting, d-f, j-l and p-r correspond to the images shown in a-c, g-i and m-o, respectively, after deconvolution and threshold steps. Note that the original immunohistochemical findings (stained structures and tissue distribution) were preserved. Scale bar: 100 μm (a-r).

Quantitative evaluation of HMGCS1 immunostaining by using the ImageJ® software yielded the values 0.31 ± 0.32 (mean percentage value ± standard deviation), 3.33 ± 1.08, and 3.86 ± 2.02 for controls, FCD IIa and FCD IIb samples, respectively. Significantly higher expressions were found in FCD IIa and FCD IIb compared to controls (p=0.02 and 0.002, respectively). No difference was found after comparing FCD IIa and FCD IIb groups (p=0.99) (Figs. 5 and 6).

**Fig. 6.**
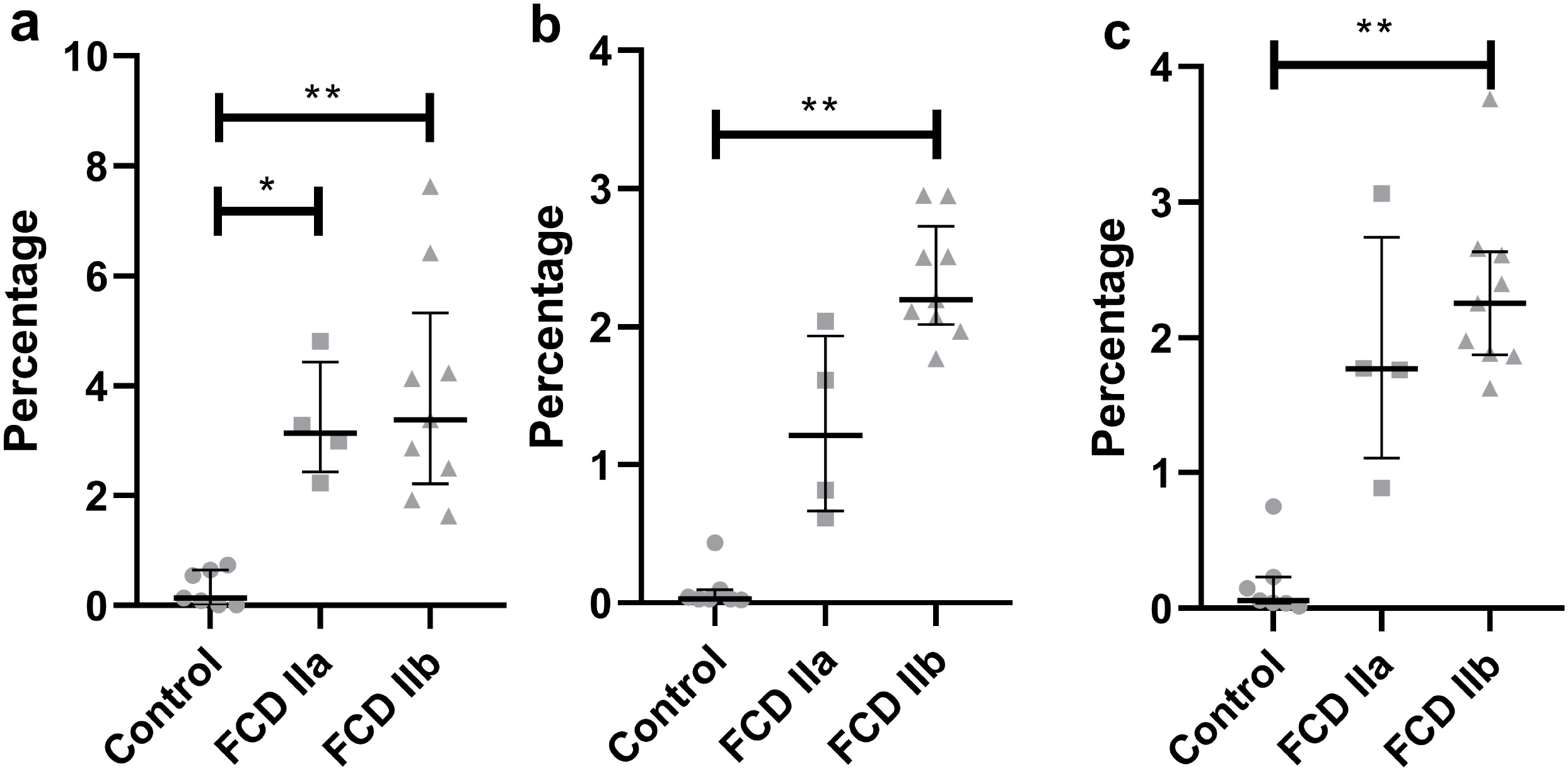
Quantification of the immunohistochemical expressions (percentage of positive pixels per total number of pixels) of HMGCS1 (a), HMGCR (b) and SQLE (c) in the gray matter of focal cortical dysplasia and controls. The graphs show scatterplots with superimposed median and interquartile range (25-75). (*) p<0.05; (**) p<0.01 (Kruskall-Wallis test followed by Dunn’s post-hoc test).

As regards HMGCR, the values for controls, FCD IIa and FCD IIb were 0.10±0.15, 1.27±0.67, and 2.34±0.42, respectively. A higher expression was observed in FCD IIb when compared to controls (p=0.0002). No differences were found when comparing controls and FCD IIa (p= 0.31), as well as FCD IIa and IIb groups (p=0.31) (Figs. 5 and 6).

Values for SQLE immunostaining were 0.18±0.26, 1.87±0.90 and 2.34±0.64 for controls, FCD IIa and FCD IIb groups, respectively. A higher expression of SQLE was found when comparing FCD IIb with controls (p=0.0008). However, no differences were found between FCD IIa and controls (p=0.09), as well as between FCD IIa and IIb (p=0.99) (Figs. 5 and 6).

Taken together, immunohistochemical detections of HMGCS1, HMGCR and SQLE paralleled most of the upregulation of the corresponding transcripts in samples with FCD, as compared to controls.

#### 3.4.2. GPNMB

Immunopositivity for GPNMB was noted only in balloon cells. Most of them showed diffuse cytoplasmic staining, which varied from light to dense. Some balloon cells, however, were immunonegative. Normal and dysmorphic neurons showed no reactivity. Similarly, other normal cell types (glia, endothelial and leptomeningeal) were negative for this marker. Since immunohistochemical reactions were performed in sections containing both gray and white matters, it was possible to identify immunopositive balloon cells in both compartments, with a random distribution but predominantly in subpial and superficial white matter regions (Fig. 7).

**Fig. 7.**
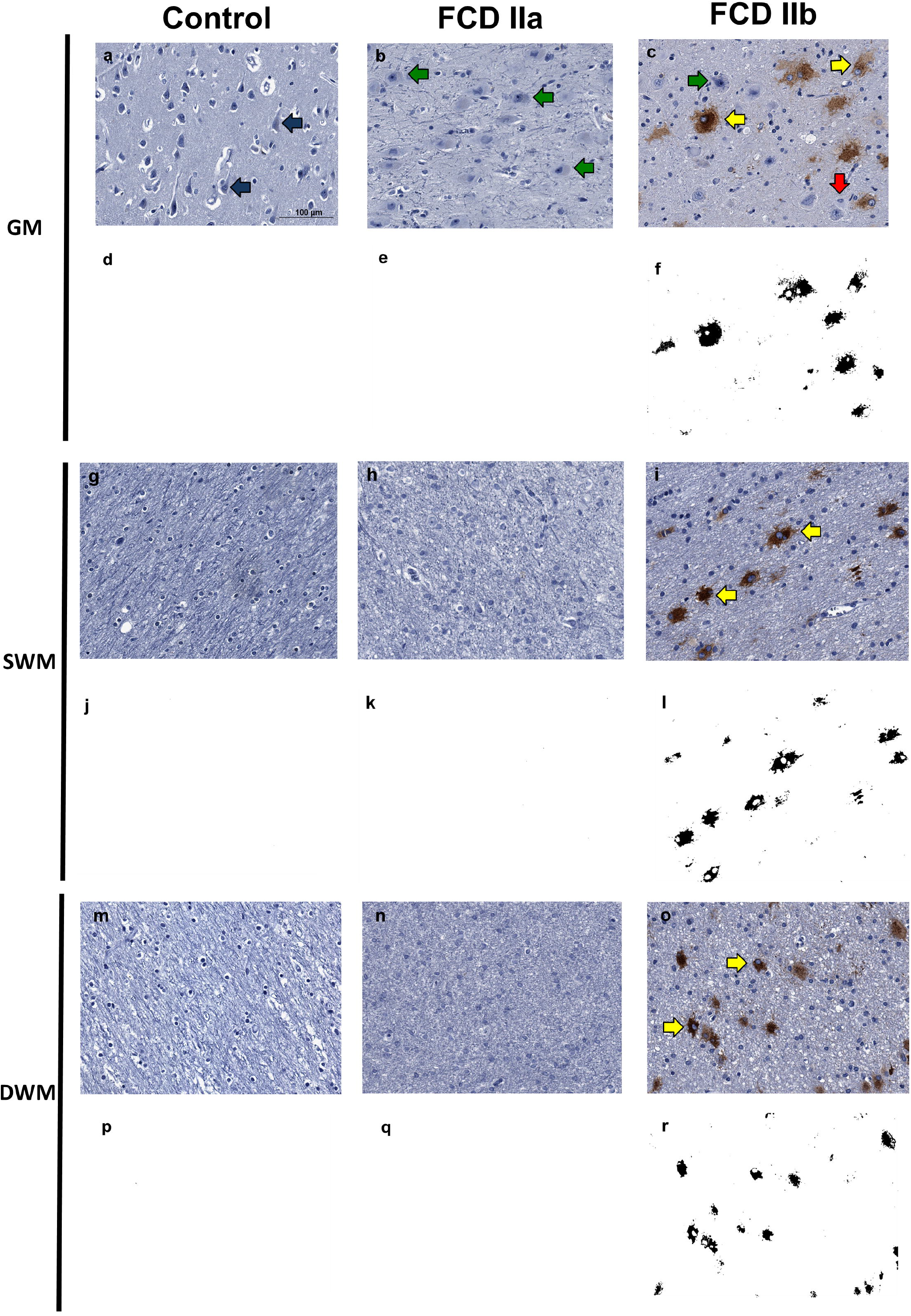
Immunohistochemical reaction for GPNMB in control, focal cortical dysplasia (FCD) ILAE Type IIa (FCD IIa) and FCD ILAE Type IIb (FCD IIb) specimens. The gray matter (GM), superficial white matter (SWM) and deep white matter (DWM) are shown in a-f, g-l and m-r, respectively. Immunopositivity was noted virtually only in balloon cells. Particularly, the staining in such cells varied from null (red arrows in c) to intense (yellow arrows in c, i, o). Both dysmorphic (green arrows in b and c) and normal (blue arrows in a) neurons were negative. Digital images of the presented immunohistochemical reactions were obtained by using the ImageJ® software (d-f, j-l, p-r). In this setting, d-f, j-l and p-r correspond to the images shown in a-c, g-i and m-o, respectively, after deconvolution and threshold steps. Note that the original immunohistochemical findings (stained structures and tissue distribution) were preserved. Scale bar: 100 μm (a-r).

After quantitative assessment of GPNMB positivity by using the ImageJ® software, the following values were verified in the superficial white matter for controls, FCD IIa and FCD IIb groups: 0.002 ± 0.001, 0.003 ± 0.001, and 1.510 ± 0.810, respectively. As regards the deep white matter, the values for controls, FCD IIa and FCD IIb were: 0.002 ± 0.001, 0.002 ± 0.001, and 1.07 ± 0.801, respectively. When considering the superficial white matter, significantly higher expression was found in FCD IIb compared to FCD IIa and controls, (p=0.02 and p=0.001, respectively). No difference was found between FCD IIa and controls (p=0.99). At the deep white matter level, no significant differences were found between the groups (FCD IIa *vs* controls: p=0.99; FCD IIb *vs* controls: p=0.07; FCD IIa vs FCD IIb: p=0.09) (Fig. 8).

**Fig. 8.**
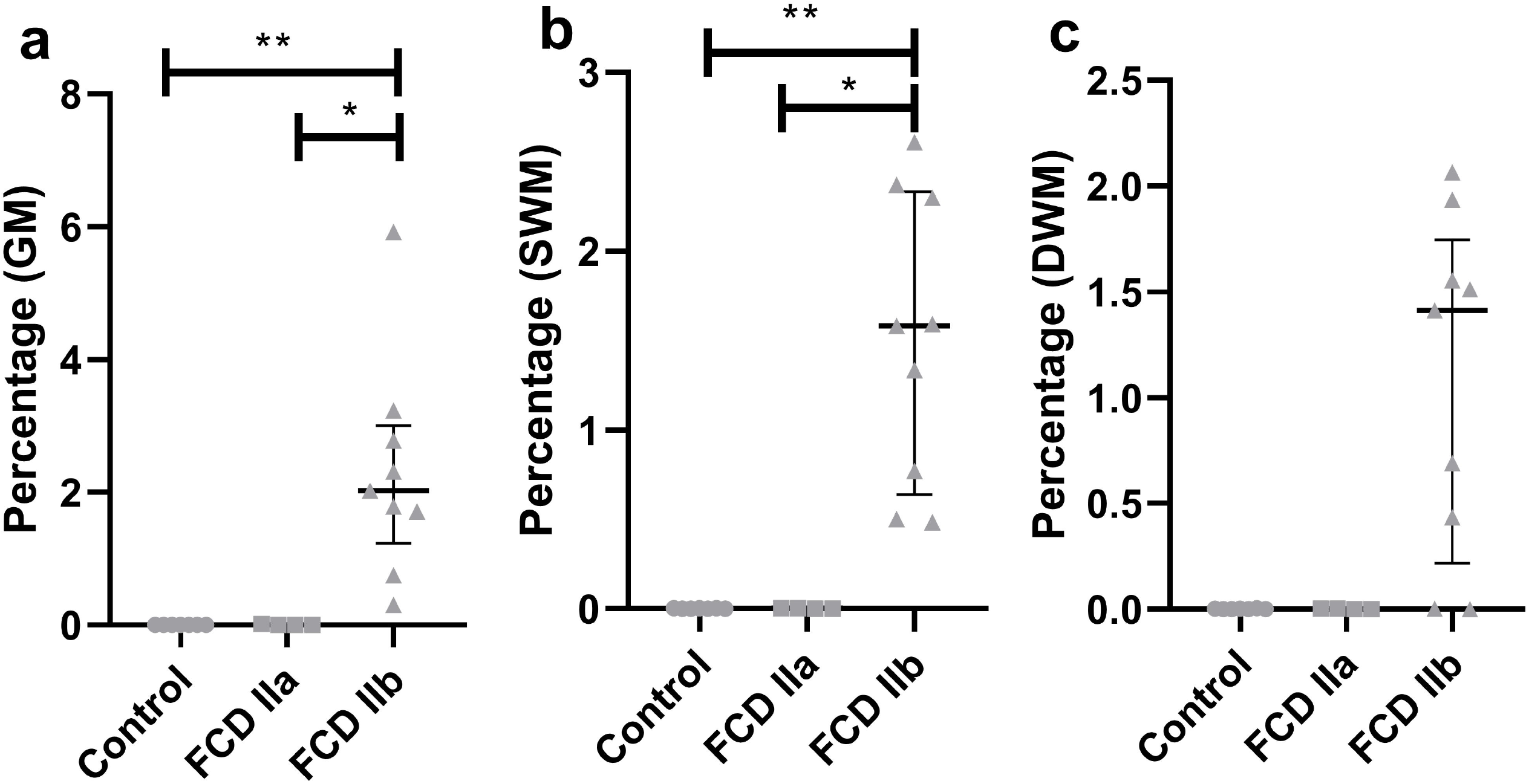
Quantification of the immunohistochemical expression (percentage of positive pixels per total number of pixels) of GPNMB in the gray matter (GM; a), superficial white matter (SWM; b) and deep white matter (DWM; c) of focal cortical dysplasia and controls. The graphs show scatterplots with superimposed median and interquartile range (25-75). (*) p<0.05; (**) p<0.01 (Kruskall-Wallis test followed by Dunn’s post-hoc test).

The observation of GPNMB-positive balloon cells in the dysplastic cortical layer prompted us to look for the corresponding gene expression in the gray matter transcriptome of each group. Likewise, *GPNMB* was found to be significantly upregulated in FCD IIb *vs* controls (Fold change = 1.08, adjusted p-value = 0.01) and downregulated in FCD IIa *vs* FCD IIb (Fold change = -1.18, adjusted p-value = 0.04). *GPNMB* was not present in the list of differentially expressed genes, when FCD IIa and control groups were compared (Figs. 7 and 8).

Quantitative evaluation of GPNMB immunostaining showed the values 0.002 ± 0.001, 0.005 ± 0.005, and 2.314 ± 1.632 for controls, FCD IIa and FCD IIb samples, respectively. A higher expression was observed in FCD IIb when compared to controls and FCD IIa (p=0.002 and p=0.01, respectively). No difference was found when comparing controls and FCD IIa (p= 0.99) (Figs. 7 and 8).

## 4. Discussion

In this work, we aimed to generate additional insights into the molecular landscape of FCD tissues by assessing the transcriptome profile, enriched cellular pathways, differential gene expression, and protein synthesis in surgical samples from patients with FCD type II.

For this purpose, we designed an original protocol to dissect the gray and white matter and evaluate each transcriptome by using a high-throughput sequencing method (RNA-Seq). Such design aimed to avoid biases due to analyses performed with tissue homogenates from both regions of the same sample (22). Overall, our results revealed different transcriptomic signatures in FCD type IIa and IIb lesions. Of note, the RNA-Seq method allowed us to obtain data not considered in previous studies focusing only on histopathological analyses of human FCD samples. Thus, such genetic data prompted us to further histopathologically and specifically investigate the expressions of cholesterol biosynthesis enzymes and GPNMB, which are respectively potential neuropathological biomarkers of a dysplastic brain cortex chronically exposed to seizures and of balloon cells.

Regarding the gray matter, we found enrichment of a wide range of cellular pathways and the differential expression of 342 and 399 genes respectively in IIa and IIb groups compared to controls. Therefore, we focused our discussion on genes not previously described in human FCDs. In this sense, both FCD type II lesions showed enrichment of the Cholesterol biosynthesis pathway and increased expression of *HMGCS1, HMGCR*, and *SQLE*. Due to the lack of previous investigations on those genes, we decided to validate our transcriptome results by performing immunohistochemical analyses of the codified proteins.

Cholesterol is a molecule with a wide range of functions, including cellular signaling and stability, fluidity, and plasma membrane permeability. Cholesterol may be synthesized by either neurons or glial cells in the central nervous system. Briefly, the initial steps involve 3-Hydroxy-3-Methylglutaryl-CoA Synthase 1 (HMGCS1) conjugating acetyl-CoA with acetoacetyl-CoA to synthesize 3-Hydroxy-3-Methylglutaryl-CoA (HMG-CoA), which is the substrate for HMG-CoA Reductase (HMGCR) to produce mevalonate. Subsequently, mevalonate is converted to squalene by Squalene Epoxidase (SQLE). In neurons, cholesterol takes part in synaptogenesis and growth of dendrites and axons (23–25).

Here, the immunohistochemical analyses of HMGCS1, HMGCR, and SQLE supported most of the upregulation of transcripts detected in FCD types IIa and IIb, compared to controls. Particularly, we observed a higher percentage of immunopositivity for the three enzymes in IIb lesions and HMGCS1 in IIa samples. The fact that the immunohistochemical expression of the downstream enzymes HMGCR and SQLE did not parallel the gene expression in IIa lesions may be due to molecular events associated with transcription and translation processes, including abundances and degradation rates of mRNA and protein, alternative splicing and post-translation changes (26–28). However, such cellular processes did not seem to be discordant regarding the initial steps of cholesterol synthesis in FCD IIa, since HMGCS1 immunoexpression was also higher in this lesion than in controls.

We, therefore, put forward the hypothesis that the enrichment of the cholesterol pathway in the context of chronic seizures due to FCD may be a tissue response to the imbalance in processes such as neurotransmission, synaptic vesicle exocytosis, and maintenance of dendritic spines and axons. Furthermore, since cytotoxic-T cell infiltrates may be observed in tissue from patients with FCD type II (29,30), cholesterol pathway enrichment may be a response to minimize resident cell damage. Indeed, cholesterol-containing lipoproteins secreted by glia cells exert anti-inflammatory and anti-apoptotic actions on glia and neurons, respectively (24,31).

Furthermore, we found that *MTRNR2L12* was the only upregulated transcript in the gray matter of FCD type IIa lesions compared to IIb. To the best of our knowledge, there are no previous reports about *MTRNR2L12* expression in surgical samples from patients with FCD. However, other authors have addressed studies on *MTRNR2* (also known as humanin) and demonstrated that the expression of the corresponding protein in human neurons (32) was associated with protective effects, such as anti-apoptotic by inhibiting BAX translocation from the cytosol to mitochondria (33), antioxidant by increasing superoxide dismutase activity (34), and preservation of synaptic connectivity by preventing dendritic atrophy caused by glutamate (35). Thus, should *MTRNR2L12* exert similar functions to those described for *MTRNR2*, it would be possible to consider a neuroprotective role for the presently detected transcript in an abnormal cortical circuitry with dysmorphic neurons, no balloon cells and chronically exposed to seizures. Future approaches could also investigate the importance of *MTRNR2L12* as a diagnostic neuropathological marker of FCD type IIa.

The analysis of the white matter transcriptome of FCD type II lesions yielded no results that we could straightforwardly associate with FCD or other brain malformations causing epilepsy. However, a review of each of the differentially expressed genes showed that *GPNMB* expression was significantly higher in the FCD IIb group than in FCD IIa and controls. Notably, such gene codifies the glycoprotein nonmetastatic melanoma protein B (GPNMB), a molecule that plays a role in motility processes, such as invasion and metastasis, in poorly metastatic melanoma cells (36). Therefore, we hypothesized that this protein could be relevant to the pathophysiology of FCD, a malformation in which abnormal cell migration is also involved. Interestingly, the additional immunohistochemical investigation of GPNMB in the current brain samples highlighted positivity virtually only in balloon cells.

As far as we know, there are no previous reports on GPNMB expression in FCD surgical samples or experimental models. On the other hand, such protein has been described to have neuroprotective, reparative, or anti-inflammatory actions in neurodegenerative disorders (37–39) and cerebral ischemia-reperfusion injury (40) Under these circumstances, GPNMB was found to be synthesized by neurons, astrocytes, and microglia in both human and murine tissues. However, the corresponding pathophysiology mechanisms are still debatable. Conversely, in our study GPNMB was only observed in balloon cells. Such discrepancy with the mentioned reports could be due to technical matters, including primary antibody clonality. Irrespective of tissue distribution differences between our study and those from others, it is conceivable that GPNMB expression by balloon cells might be associated with neuroprotection, cellular reparation, and anti-inflammatory action. As proposed above, FCD type II may involve disruptions to neurotransmission and maintenance of neuronal membrane structures and tissue inflammation (29,30).

A relevant finding of the current investigation is GPNMB as a potential diagnostic neuropathological biomarker of balloon cells. In fact, we did not identify other cell types (normal or abnormal) immunopositive for this molecule by using the primary antibody, whose clone is SP299. Future studies with larger cohorts and different epilepsy centers are necessary to validate such findings and support their use in the diagnostic routine. A potential contribution would be to characterize better small surgical samples from individuals with clinical neuroimaging diagnosis of FCD type IIb but whose neuropathological analysis only shows dysmorphic neurons. In this context, the addition of anti-GPNMB antibody to the currently recommended immunohistochemical markers by the ILAE (13) could help in the search for balloon cells, thus, improving the histopathological work-up and classification of the dysplastic cortex.

Somatic mutations in genes belonging to the mTOR pathway have been identified in tissue from patients with FCD type IIb (11). Among the tissue samples studied in the present work, we found two with somatic missense variants in the *MTOR* gene. One is a variant previously linked to FCD type IIb (c.5930C>G, p.Thr1977Arg, (21), the second is a novel variant classified as VUS (c.4375G>C, p.Ala1459Pro). Although we cannot unequivocally conclude that the c.4375G>C may be associated with FCD type IIb, we believe it is most likely pathogenic since it has never been reported in control databases, including in an ethnically matched database (www.bipmed.org; (19)). In addition, the in-silico prediction on protein function indicates that the variant is probably/possible pathogenic by three of the four algorithms used.

In conclusion, our study provides novel and high-quality data about gene expression and enriched cellular pathways obtained from surgical samples of patients with FCD type IIa and IIb. The current genetic and histopathological results, suggesting cellular events associated with neuroprotection, may base future clinical and experimental approaches to further understand the pathophysiological events associated with FCDs and tissue response to seizures. Furthermore, upregulations of *MTRNR2L12* in FCD IIa and GPNMB in FCD IIb might be potential neuropathological biomarkers of a brain cortex chronically exposed to seizures and for balloon cells, respectively. Our results highlighted potential pathophysiological mechanisms and contributed to tissue characterization of a frequent cause of focal epilepsy.

## Data Availability

All data produced in the present study are available upon reasonable request to the authors.

## Conflict of interest disclosure

The authors declare that they have no conflict of interest.

## Ethics Approval and Consent to Participate

This study was performed according to Brazilian and German institutional ethical approvals (UNICAMP - CEP#470/2003 and CAAE: 12112913.3.0000.5404; FAU - FP7 health program, DESIRE grand agreement#602531). Written informed consent was obtained from all participants.

## Author contribution statement

FR, IL-C, FC, ASV, and IB contributed to the study design. GRAM, MCPA, JVGT, AS, ASV, GGZ, PAORAA, EG, HT, MKMA, VSA, WS, CLY, RC, IB, FC, IL-C, and FR obtained and analyzed the data. GRAM, JVGT, MCPA, ASV, IB, FC, IL-C, and FR drafted the manuscript and figures. All authors reviewed and approved the final version of the manuscript.

## Acknowledgements

This study was sponsored by grants from Fundação de Amparo à Pesquisa do Estado de São Paulo (FAPESP; 2013/07559-3, 2016/50486-5 and 2019/08259-0) and FAEPEX UNICAMP (2037/19, 2428/20 and 2059/22).

## Supplementary Material

Transcriptome data (enriched pathways and gene expression) of focal cortical dysplasia (FCD) ILAE type IIa (FCD IIa) and FCD ILAE type IIb (FCD IIb) samples. Data of the gray and white matters are presented separately. Datasheets 1 and 2 show the enriched pathways, p-values and corresponding differentially expressed genes of the gray matter of FCD IIa and IIb groups, respectively, versus controls (autopsy). Datasheets 3 – 8 present gene symbols (or Ensembl code when gene symbol was not available), Fold Change (log_2_fold change) values relative to the groups indicated in each datasheet designation, p-values and adjusted p-values. Ctrl: control; GM: gray matter (cortical layer); WM: white matter.

## Notes

### Competing Interest Statement

The authors have declared no competing interest.

### Funding Statement

This study was sponsored by grants from Fundacao de Amparo a Pesquisa do Estado de Sao Paulo (FAPESP; 2013/07559-3, 2016/50486-5 and 2019/08259-0) and FAEPEX UNICAMP (2037/19, 2428/20 and 2059/22).

### Author Declarations

Ethics committee/IRB of the University of Campinas (UNICAMP; Campinas, Brazil) and the University Hospital Erlangen (FAU; Erlangen, Germany) gave ethical approval for this work (UNICAMP - CEP#470/2003 and CAAE: 12112913.3.0000.5404; FAU - FP7 health program, DESIRE grand agreement#602531)

