## Supplementary material for "Transcriptome analyses of the cortex and white matter of focal cortical dysplasia type II: insights into disease mechanisms and tissue characterization": Table 1

**Table 1 – Clinical data of the individuals with diagnosis of FCD ILAE Type II, whose surgical samples were** **submitted for transcriptome analysis and/or immunohistochemical validation.**

| **Group** | **#** | **Gd** | **Age at onset (range in years)** | **Age at surgery (range in years)** | **Duration of crises (range in years)** | **Lobe / side** | **Transcriptome** | | **IHC** | |
| --- | --- | --- | --- | --- | --- | --- | --- | --- | --- | --- |
|  |  |  |  |  |  |  | **G** | **W** | **G** | **W** |
| FCD IIa | 1 | F | 6-10 | 11-15 | 0-5 | Fr - R |  |  |  |  |
|  | 2 | F | 0-5 | 11-15 | 11-15 | Fr - R |  |  |  |  |
|  | 3 | F | 6-10 | 31-35 | 21-25 | Fr - R |  |  |  |  |
|  | 4 | M | 0-5 | 16-20 | 11-15 | Fr - R |  |  |  |  |
|  | 5 | M | 0-5 | 6-10 | 0-5 | Fr - L |  |  |  |  |
|  | 6 | F | 0-5 | 46-50 | 46-50 | Fr - L |  |  |  |  |
|  | 7 | M | 6-10 | 21-15 | 16-20 | Fr - L |  |  |  |  |
|  | 8 | F | 0-5 | 16-20 | 16-20 | Fr - L |  |  |  |  |
|  | 9 | F | 0-5 | 6-10 | 6-10 | Fr - L |  |  |  |  |
|  | 10 | F | 0-5 | 11-15 | 11-15 | Fr - R |  |  |  |  |
|  | 11 | F | 6-10 | 31-35 | 26-30 | Fr - R |  |  |  |  |
| FCD IIb | 12 | M | 0-5 | 21-25 | 21-25 | Fr - L |  |  |  |  |
|  | 13 | F | 11-15 | 21-25 | 6-10 | Fr - L |  |  |  |  |
|  | 14 | F | 0-5 | 0-5 | 0-5 | Fr - L |  |  |  |  |
|  | 15 | F | 0-5 | 11-15 | 11-15 | Fr - R |  |  |  |  |
|  | 16 | F | 0-5 | 6-10 | 0-5 | Fr - R |  |  |  |  |
|  | 17 | F | 11-15 | 41-45 | 26-30 | Fr - L |  |  |  |  |
|  | 18 | F | 11-15 | 31-35 | 16-20 | Fr - L |  |  |  |  |
|  | 19 | F | 11-15 | 41-45 | 26-30 | Fr - R |  |  |  |  |
|  | 20 | M | 0-5 | 16-20 | 17-20 | Fr - R |  |  |  |  |
|  | 21 | M | 6-10 | 41-45 | 6-10 | Pr - R |  |  |  |  |
|  | 22 | F | 0-5 | 31-35; 41-45 | 26-30;  36-40 | T - R |  |  |  |  |
|  | 23 | M | 6-10 | 36-40 | 31-35 | Fr - R |  |  |  |  |
|  | 24 | F | 0-5 | 26-30 | 21-25 | Pr - R |  |  |  |  |
|  | 25 | F | 16-20 | 30-35 | 11-15 | O - L |  |  |  |  |

All samples submitted for transcriptome analysis were obtained from the frontal lobe, which was defined as the epileptogenic zone after clinical and neuroimaging investigation. Each rectangle colored in gray in the “Transcriptome” and “IHC” columns indicates the technique performed with the corresponding sample from the same individual. For transcriptome analyses, nine (n=9; individuals #1-9) and 8 (n=8; individuals #12-19) specimens were obtained for FCD IIa and IIb, respectively. Regarding gray matter investigation, seven (n=7) specimens were used for FCD IIa and six (n=6) for IIb. Assessment of the white matter was performed in seven (n=7) samples of each FCD II type. Please note that, dissected samples from individuals #2, #5, #7, #12, #16, and #18 could only be used for either gray other white matter analysis due to available amount or technical issues (alignment ratio of the generated sequences with the reference genome was less than 80% and 50% for gray and white matter, respectively). Materials from individuals #10, #11 and #20-25 were used for immunostainings only. F: female; FCD: focal cortical dysplasia; FCD IIa: FCD International League Against Epilepsy (ILAE) Type IIa; FCD IIb: FCD International League Against Epilepsy (ILAE) Type IIb; Fr: frontal; G: gray matter (cortical layer); IHC: immunohistochemistry; Gd: gender; L: left; M: male; O: occipital; Pr: parietal; R: right; T: temporal; W: white matter; #: number of the individual in the present study.
