## Supplementary material for "Transcriptome analyses of the cortex and white matter of focal cortical dysplasia type II: insights into disease mechanisms and tissue characterization": Table 2

**Table 2 – Clinical data of the controls, whose samples were submitted for transcriptome analysis and/or immunohistochemical validation.**

| **Group** | **#** | **Gd** | **Age at death**  **(range in years)** | **Lobe** | **Transcriptome** | | **IHC** | |
| --- | --- | --- | --- | --- | --- | --- | --- | --- |
|  |  |  |  |  | **G** | **W** | **G** | **W** |
| Control | 26 | M | 50-55 | Fr |  |  |  |  |
|  | 27 | F | 36-40 | Fr |  |  |  |  |
|  | 28 | F | 41-45 | Fr |  |  |  |  |
|  | 29 | M | 66-70 | Fr |  |  |  |  |
|  | 30 | F | 31-35 | Fr |  |  |  |  |
|  | 31 | M | 56-60 | Fr |  |  |  |  |
|  | 32 | M | 61-65 | Fr |  |  |  |  |
|  | 33 | F | 31-35 | Fr |  |  |  |  |

All samples were obtained from the frontal lobe. Gross and microscopic evaluations showed no alteration in both gray and white matters. Each rectangle colored in gray in the “Transcriptome” and “IHC” columns indicates the technique performed with the corresponding sample from the same individual. Samples from individuals #26-29 were used for transcriptome (n=4) investigations of the gray and white matters. Moreover, samples from individuals #27-33 were submitted to immunostaining. Thus, seven (n=7) specimens were used for gray and white matter analyses. F: female; Fr: frontal; G: gray matter (cortical layer); IHC: immunohistochemistry; Gd: gender; M: male; W: white matter; #: number of the individual in the present study.
